# Public acceptability, bioethics, and legal governance of wastewater and environmental surveillance for measles in South Africa

**DOI:** 10.64898/2026.09.04.26362243

**Authors:** Rukudzo Matanda, Rochelle H. Holm, Sibonginkosi Maposa, Lebohang Rabotapi, Mukhlid Yousif, Kerrigan McCarthy

## Abstract

**Objectives:** This study investigates public opinion regarding the use of wastewater and environmental surveillance for measles in South African urban centres across ethical, legal, and public health policy themes, and maps these opinions onto existing bioethical and legal precedents.

**Design:** In-person field survey.

**Setting:** Three metropolitan catchment areas in Gauteng Province, South Africa (City of Tshwane, City of Ekurhuleni, and City of Johannesburg).

**Participants:** A sample of 100 adult community members residing or working within the selected sewer catchment areas.

**Primary outcome measures:** Baseline wastewater and environmental surveillance awareness, target-specific acceptability, spatial monitoring preferences, data-sharing comfort, and legal/privacy perceptions.

**Results:** Baseline awareness of WES was low (19%). Support was high for monitoring a range of biological and chemical targets using wastewater and environmental surveillance. However, acceptability was dependent on spatial scale: 83% supported macro-level "city-wide" monitoring, whereas support fell to 28% for localized sampling (individual homes and schools). Institutional trust in data handling was high among scientific and public health bodies.

**Conclusions:** Public acceptability of WES for communicable diseases, particularly measles, in urban South Africa is high but conditional upon macro-level data aggregation ("anonymity by aggregation") and governance by dedicated scientific entities. Given strong community support for measles surveillance and its alignment with the country’s bioethical and legal frameworks, the bioethical and legal requirements for this surveillance approach can be satisfied, paving the way for routine implementation of wastewater and environmental surveillance for communicable diseases.

## INTRODUCTION

Wastewater and environmental surveillance (WES) is recognized globally as a cost-effective, non-invasive tool for monitoring population-level public health threats [1]. In low- and middle-income countries, such as South Africa, where limited resources, underreporting, and delays in healthcare-seeking often undermine the effectiveness of traditional clinical surveillance, WES offers a cost-effective, non-invasive, population-level early warning system [2–5]. However, passive biological monitoring raises distinct bioethical and legal challenges regarding autonomy, privacy, non-consensual data collection, and governance [6,7]. As the field continues to expand, low- and middle-income countries need to better understand community acceptability and privacy concerns surrounding WES for disease monitoring and how these considerations map onto bioethics and legal governance frameworks; herein, we focus on WES for measles in South Africa.

Measles is a highly contagious, vaccine-preventable, viral disease. Global elimination strategies, guided by the WHO, emphasize closing immunity gaps and developing novel surveillance tools [8, 9]. South Africa experiences recurrent, large-scale, measles outbreaks. Despite a routine two-dose Expanded Programme on Immunisation schedule administered at 6 and 12 months, routine coverage remains insufficient to maintain the 95% population-immunity threshold [9–12].

Traditional measles clinical surveillance in South Africa relies on the Notifiable Medical Conditions Surveillance System (NMCSS) [11]. Under this framework, clinicians identify suspected measles cases using standard clinical definitions of fever and rash combined with cough, coryza, or conjunctivitis, and collect blood samples for laboratory IgM testing. However, this model has critical operational limitations: mild or subclinical cases may bypass clinical detection entirely, leaving transmission chains undetected, and reporting delays may further impede timely public health response.

The concept of WES first emerged in the 1940s and involves the collection, detection, and analysis of untreated municipal wastewater for selected biological or chemical markers [1]. In disease monitoring applications, WES potentially captures pathogen shedding from both symptomatic and asymptomatic individuals across entire catchment areas [1]. In South Africa, WES studies have shown that SARS-CoV-2 signal concentrations in municipal wastewater strongly correlate with clinical caseloads while uncovering hidden infection hotspots in low-income informal settlements [3, 4]. Measles WES is also established, as the measles virus is excreted in urine for extended periods [13, 14]; WES in South Africa has been used to detect measles virus in raw wastewater samples [2, 5].

Traditional methods for public health surveillance operate under a bioethical framework where individual autonomy is balanced against the collective duty to prevent disease [1, 15, 16]. Non-consensual surveillance is ethically defensible provided proportionality, risk minimization, and public feedback are maintained [16]. The ethical deployment of WES in South Africa is anchored in constitutional, statutory, and judicial frameworks. Section 14 of the South African Constitution guarantees the right to privacy, while Section 27 obligates the State to take reasonable measures to protect public health [17]. At the statutory level, the Protection of Personal Information Act (POPIA) explicitly exempts fully anonymized or aggregate health data from individual consent mandates [18], which may be interpreted to allow macro-level municipal WES, whereas high-resolution targeting at the household level may breach statutory protection. Furthermore, the National Health Act [19] and Disaster Management Act [20] legally empower health authorities to conduct non-consensual disease tracking during public health emergencies or major disease threats, but are less clear on routine WES as an early warning surveillance system.

While legal mechanisms allow for non-consensual WES, WES still depends heavily on public trust and alignment with community privacy expectations. This study investigates public opinion regarding the use of WES for measles in South African urban centres across ethical, legal, and public health policy themes, and maps these opinions onto existing bioethical and legal precedents.

## METHODS

### Study design and setting

This study utilized a cross-sectional field evaluation design to assess public knowledge, attitudes, and practices (KAP) regarding the ethics, privacy, and acceptability implications of WES with a focus on measles. The study was nested within the established National Institute for Communicable Diseases (NICD) surveillance network in Gauteng Province, South Africa. Data collection occurred across three well-characterized wastewater catchment areas [21]:

1. City of Tshwane (CoT): Daspoort catchment (Laudium and Atteridgeville; population ∼83,500; >98% flush toilet connectivity).
2. City of Ekurhuleni (CoE): Olifantsfontein catchment (Thembisa; population ∼463,109; 84.7% sewer connectivity).
3. City of Johannesburg (CoJ): Bushkoppies catchment (Klipspruit, Soweto; population ∼16,923; 98.4% sewer connectivity).

### Participants and recruitment

A convenience sample of 100 participants was recruited during routine wastewater sampling between 5–27 November 2025. Field researchers accompanied NICD WES teams to routine collection points. Community members present at or near these sample points were invited to participate. Eligible participants were aged 18 years or older, resided or worked within the target municipalities, were able to complete the questionnaire in English, and provided informed written consent. Exclusions included individuals aged under 18 years, individuals who declined consent, and repeat respondents.

### Survey instrument and data collection

Data were collected via in-person, paper-based questionnaires adapted from a validated tool used in Malawi by Jeboda et al. [22] (see Supplementary Material for full survey instrument). Prior to administration, a brief, standardized verbal explanation of WES mechanics was delivered to resolve potential conflation between sewage surveillance and drinking water purification. This clarification was considered necessary to promote a consistent understanding of WES among respondents and to support informed responses to questions regarding its acceptability and potential applications. The questionnaire gathered data on demographic characteristics, baseline WES awareness, potential wastewater detection targets and respondent comfort levels, spatial sampling preferences, and privacy boundaries using single-choice, multiple-response, and 5-point Likert scales.

### Data management and analysis

Responses were entered into a REDCap database and analysed using R software. Likert scale responses were summarized using descriptive statistics (frequencies, percentages, modes). Contradictory responses regarding consent dynamics were retained to reflect public conceptual nuances regarding non-consensual monitoring.

Comparative case law was identified through dedicated legal databases, specifically the Southern African Legal Information Institute (SAFLII), as well as LexisNexis. Relevant statutes, policies, and ethical codes were identified through targeted searches of official government and public health websites. The review examined constitutional rights to privacy and health; key legislation, including the POPIA, the National Health Act [19], and the Disaster Management Act [20]; and Health Professions Council of South Africa (HPCSA) guidelines [23].

### Study ethics approval, consent, and voluntary participation

The study protocol was approved by the Human Research Ethics Committee (HREC) of the University of the Witwatersrand (Clearance Certificate No. M250305). All participants provided written informed consent. Participation was voluntary, and data were anonymised using unique numerical identifiers to minimize potential response bias introduced by field apparel.

## RESULTS

A total of 100 adult community members across three major metropolitan municipalities in Gauteng Province participated in the study: the CoE (79%, n = 79), CoJ (15%, n = 15), and CoT (6%, n = 6). Baseline public awareness of WES was low across the overall cohort. Only 19% of respondents reported prior knowledge or familiarity with WES disease surveillance, such as determining whether measles is present in the community.

### WES scope acceptability

Respondents reported consistently high levels of comfort regarding the application of WES to monitor various biological and chemical targets (Table 1). The acceptability of WES as a public health tool was high (being somewhat or very comfortable with monitoring): 83% of respondents were somewhat or very comfortable with monitoring SARS-CoV-2; 83% for serious diseases (e.g., tuberculosis); 82% for common infectious diseases (e.g., measles, malaria, and influenza); and 79% for sexually transmitted infections (e.g., HIV and mpox). Support was similarly robust for monitoring non-communicable and lifestyle indicators: 89% of respondents were somewhat or very comfortable with monitoring nutritional indicators; 86% for dagga, cocaine, tik, and nyaope; 84% for smoking, coffee drinking, and alcohol use; 81% for hormones indicating population happiness or stress; 84% for prescription drugs (high blood pressure and diabetes medication, and antibiotics); and 81% for traditional medicines (e.g., umhlonyane). In terms of using WES for environmental health, 84% of respondents reported being somewhat or very comfortable with monitoring for harmful substances such as pesticides (e.g., organophosphates and rat poison), and 86% were somewhat or very comfortable with monitoring for harmful substances such as mining chemicals or air pollutants. Across all evaluated targets, the proportion of respondents reporting discomfort (very or somewhat uncomfortable) remained below 10%.

**Table 1.** Participant comfort levels with wastewater and environmental surveillance of biological and chemical targets. 5–27 November 2025, South Africa. N = 100.

| **Target** | **% of respondents in each category** |  |  |  |  |
| --- | --- | --- | --- | --- | --- |
|  | **1**<br>**(very**<br>**uncomfortable)** | **2** | **3** | **4** | **5**<br>**(very**<br>**comfortable)** |
| **SARS-CoV-2** | 6% | 1% | 10% | 4% | 79% |
| **Common infectious diseases** | 7% | 1% | 10% | 2% | 80% |
| **Serious diseases** | 6% | 3% | 8% | 3% | 80% |
| **Sexually transmitted infections**<br>**(e.g., HIV and mpox)** | 8% | 3% | 10% | 3% | 76% |
| **Markers of good nutrition** | 4% | 1% | 6% | 7% | 82% |
| **Hormones that show how happy**<br>**a population is** | 6% | 2% | 11% | 3% | 78% |
| **Prescription drugs** | 5% | 1% | 10% | 4% | 80% |
| **Traditional medicine** | 9% | 1% | 9% | 5% | 76% |
| **Smoking, coffee drinking** | 5% | 1% | 10% | 6% | 78% |
| **Dagga, nyaope, cocaine** | 6% | 1% | 7% | 5% | 81% |
| **Harmful pesticides (e.g., rat**<br>**poison)** | 8% | 1% | 7% | 6% | 78% |
| **Mining chemicals** | 5% | 2% | 7% | 6% | 80% |

### WES acceptability as a complement to clinical testing

When specifically assessing the application of WES for measles surveillance, a complex dual preference emerged which may indicate that respondents did not understand the scope of WES and public health surveillance. While 73% of respondents strongly agreed that the Department of Health (DoH) should employ wastewater testing to detect circulating measles virus, 49% strongly agreed that measles WES should not replace clinical surveillance (Table 2).

**Table 2.** Participant responses regarding acceptability of WES for measles as a complement to clinical testing, 5–27 November 2025, South Africa. N = 100.

| Question | Number (%) of persons responding in each category |  |  |  |  |
| --- | --- | --- | --- | --- | --- |
|  | 1<br>(strongly disagree) | 2 | 3 | 4 | 5<br>(strongly agree) |
| DoH should use WES to find circulating measles virus | 2% | 3% | 19% | 3% | 73% |
| DoH should only use direct testing of people to find measles cases | 12% | 8% | 26% | 5% | 49% |
| DoH should ask the community's permission to test wastewater for measles | 18% | 7% | 18% | 6% | 51% |
| DoH does not need to ask everyone in the community if it is acceptable to test wastewater for measles | 18% | 5% | 18% | 3% | 56% |

### Geographic scale and acceptability of sampling contexts

When comparing preferred geographic monitoring scales, a clear dichotomy emerged. High baseline acceptability for public health targets did not translate to uniform support across all geographic categories. Regarding sampling locations, respondents expressed a strong preference for aggregated, macro-level surveillance. Broad-scale macro-surveillance targeting “the entire city” received 83% support. Acceptability dropped to 36% for monitoring specific suburbs and 33% for informal settlements, 30% for shopping areas and taxi ranks where communal toilets may be common, 28% for schools and homes, 26% for businesses and prisons, and 25% for church/mosque/synagogue.

### Privacy concerns, data sharing, and institutional trust

Participants reported higher levels of trust in health sector organizations and scientific bodies around WES for measles. Comfort levels were highest for sharing personal information and wastewater data with local Primary Health Care clinics (82% for very or somewhat comfortable), the National Department of Health (NDoH) (79% for very or somewhat comfortable), and the National Institute for Communicable Diseases (NICD) (76% for very or somewhat comfortable). Participants were also comfort sharing data with ward councillors (76% for very or somewhat comfortable).

Negative views regarding privacy intrusion were few. Only 16% of participants agreed that WES constitutes an invasion of personal privacy. Notably, a substantial level of uncertainty remained within the cohort, with 41% of respondents indicating they were unsure about the privacy implications of the technology.

### Mapping the KAP to South African statutory and regulatory frameworks

Participant views closely match South African statutory and regulatory frameworks, showing that public preferences mirror the law (Table 3). High support for city-wide testing (83%) and low support for household testing (28%) reflect rules under POPIA [18] and case law in De Jager v Netcare [24]. In *De Jager v Netcare Limited and Others*, the South African High Court established that health-related observations in private domains require strict consent, whereas data captured in public spaces carries a reduced expectation of privacy [24]. Furthermore, high trust in health agencies matches POPIA Section 32, which limits data handling to health bodies [18]. Finally, accepting outbreak tracking without consent aligns with the Disaster Management Act [20] and court rulings in Goliath [25] and Ngobeni [26], which prioritize public health safety over personal privacy during health crises. In *Minister of Health of the Western Cape v Goliath and Others* [25] and *Ngobeni v National Health Laboratory Service (NHLS) and Others* [26], the courts ruled that individual privacy rights are legally subordinate to the broader public interest during communicable disease outbreak management.

**Table 3:**
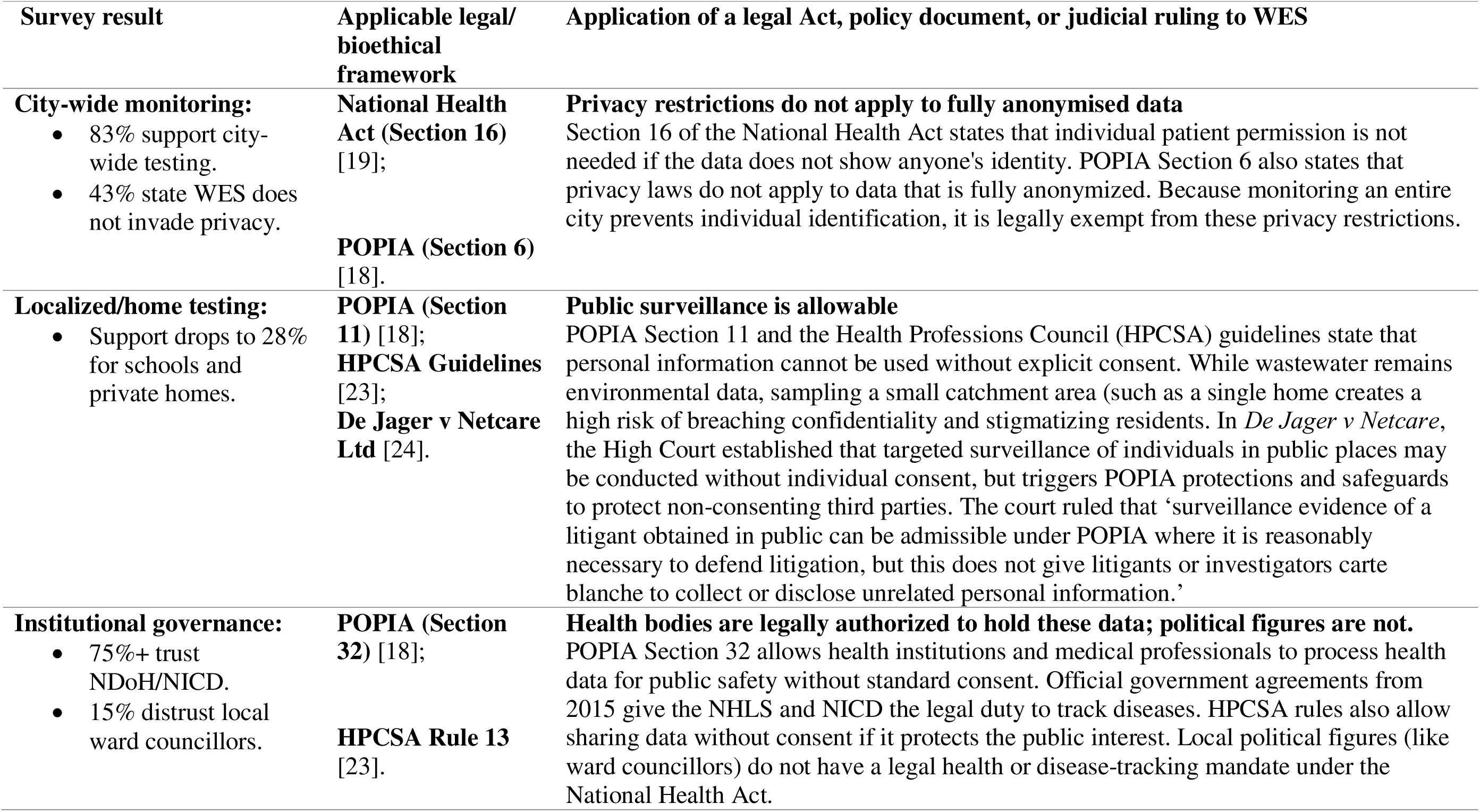
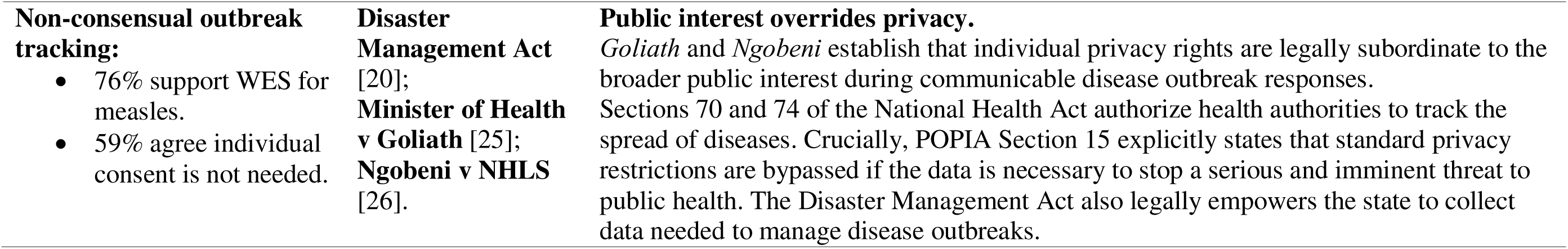
Participant knowledge, attitudes, and practices and corresponding South African statutory and regulatory frameworks.

| Survey result | Applicable legal/ bioethical framework | Application of a legal Act, policy document, or judicial ruling to WES |
| --- | --- | --- |
| **City-wide monitoring:**<br>83% support city-wide testing.<br>43% state WES does not invade privacy. | **National Health Act (Section 16)** [19];<br>**POPIA (Section 6)** [18]. | **Privacy restrictions do not apply to fully anonymised data**<br>Section 16 of the National Health Act states that individual patient permission is not needed if the data does not show anyone's identity. POPIA Section 6 also states that privacy laws do not apply to data that is fully anonymized. Because monitoring an entire city prevents individual identification, it is legally exempt from these privacy restrictions. |
| **Localized/home testing:**<br>Support drops to 28% for schools and private homes. | **POPIA (Section 11)** [18];<br>**HPCSA Guidelines** [23];<br>**De Jager v Netcare Ltd** [24]. | **Public surveillance is allowable**<br>POPIA Section 11 and the Health Professions Council (HPCSA) guidelines state that personal information cannot be used without explicit consent. While wastewater remains environmental data, sampling a small catchment area (such as a single home creates a high risk of breaching confidentiality and stigmatizing residents. In *De Jager v Netcare*, the High Court established that targeted surveillance of individuals in public places may be conducted without individual consent, but triggers POPIA protections and safeguards to protect non-consenting third parties. The court ruled that 'surveillance evidence of a litigant obtained in public can be admissible under POPIA where it is reasonably necessary to defend litigation, but this does not give litigants or investigators carte blanche to collect or disclose unrelated personal information.' |
| **Institutional governance:**<br>75%+ trust NDoH/NICD.<br>15% distrust local ward councillors. | **POPIA (Section 32)** [18];<br>**HPCSA Rule 13** [23]. | **Health bodies are legally authorized to hold these data; political figures are not.**<br>POPIA Section 32 allows health institutions and medical professionals to process health data for public safety without standard consent. Official government agreements from 2015 give the NHLS and NICD the legal duty to track diseases. HPCSA rules also allow sharing data without consent if it protects the public interest. Local political figures (like ward councillors) do not have a legal health or disease-tracking mandate under the National Health Act. |
| **Non-consensual outbreak tracking:**<br>• 76% support WES for measles.<br>• 59% agree individual consent is not needed. | **Disaster Management Act** [20];<br>**Minister of Health v Goliath** [25];<br>**Ngobeni v NHLS** [26]. | **Public interest overrides privacy.**<br>*Goliath* and *Ngobeni* establish that individual privacy rights are legally subordinate to the broader public interest during communicable disease outbreak responses.<br>Sections 70 and 74 of the National Health Act authorize health authorities to track the spread of diseases. Crucially, POPIA Section 15 explicitly states that standard privacy restrictions are bypassed if the data is necessary to stop a serious and imminent threat to public health. The Disaster Management Act also legally empowers the state to collect data needed to manage disease outbreaks. |

## Discussion

This evaluation of public KAP regarding WES in South Africa, with a focus on measles, reveals low baseline awareness of WES, paired with widespread support for large-scale city surveillance over localized household or institutional monitoring where individual identification risks are high. Beyond measles, public comfort remained high even for more sensitive or potentially stigmatizing indicators, including traditional medicine. When evaluated against South African policy and legal frameworks, our findings identified a strong alignment between public preferences and statutory privacy rules.

When comparing our findings with from Malawi [22] and the United States [27] baseline, South African respondents similarly demonstrated high general support for WES as a public health tool. The high proportion of neutral or uncertain responses in our study suggests that public hesitation is driven more by a lack of familiarity with surveillance mechanisms rather than active opposition or entrenched privacy concerns. Comparing Malawi [20] and South Africa, both countries strongly supported city-wide testing (83% in South Africa vs. 87% in Malawi), but support in South Africa was much lower for testing in schools (28% vs. 95%), homes (28% vs. 86%), prisons (26% vs. 87%), and businesses (26% vs. 79%). Consequently, future public health policies must prioritize proactive, transparent educational campaigns to bridge this knowledge gap and highlighting the need for localized, context-specific communication strategies.

Our results provide real-world evidence for the identifiability continuum discussed in global wastewater ethics [6, 7], showing also in South Africa that privacy hesitancy around WES increases as sampling areas get smaller. Seminal ethical frameworks for wastewater surveillance, such as those proposed by Canadian Water Network’s COVID-19 Wastewater

Coalition [28] and Coffman et al. [29], caution that as spatial resolution increases (i.e., smaller catchment areas), the risk of group stigmatization and individual re-identification increases. Our findings demonstrate that the South African public intuitively understands this boundary, favouring anonymity by aggregation" Residents recognize their privacy is secure when diluted within a massive urban environment, but perceive localized monitoring as a potential threat. Future South African WES frameworks must formally establish minimum spatial boundaries to align with both public preference and international bioethics standards, preventing the stigmatization of small, identifiable groups.

Participants exhibited high levels of trust in dedicated health authorities (NDoH and NICD), who would be most likely to lead WES at a local, regional, or national level for South Africa. This is despite widespread distrust of public health institutions in South Africa. As well, local ward councillors are directly responsible for delivering municipal services, leading community health education, and implementing local disease responses, all of which depend heavily on public trust to succeed. As demonstrated during the COVID-19 pandemic, major health crises require political leadership to implement broad public interventions, meaning health data cannot simply be isolated from political administrators. Rather, clear governance frameworks should exist to uphold privacy. In the South African context, where historical inequities and bureaucratic mistrust remain prevalent, this depoliticization is even more critical. Keeping WES data within scientific and medical pipelines validates the foundational goals of the proposed National Public Health Institute of South Africa (NAPHISA) framework [30]. The long-term solution is not complete political exclusion; instead, health authorities must focus on building political trustworthiness, and political leaders must develop competence in managing health issues. Training local councillors on health data privacy and clear governance boundaries will allow them to support WES initiatives effectively without eroding public trust.

Crucially, boundaries established by the community respondents mirror statutory thresholds, constitutional tensions, and recent case law within South African jurisprudence. The South African Constitution necessitates a delicate balance between the individual’s right to privacy (Section 14) and the state’s obligation to provide public health protection (Section 27) [17]. The survey data indicate that the public instinctively navigates this constitutional balance when evaluating surveillance frameworks. The public’s strong rejection of localized household testing (28%) mirrors the judicial precedent set in *De Jager v Netcare Limited and Others* [24]. In this case, the court established that while health-related observations in public domains carry a diminished expectation of privacy, capturing such data in private settings constitutes a strict privacy violation. The survey respondents intuitively applied this legal principle: once wastewater enters the public municipal domain, it is viewed as an aggregate, depersonalized asset, but at the household level, it remains legally protected private information.

Additionally, the survey confirms public alignment with statutory emergency mandates. The majority consensus (59%) that universal individual consent is unnecessary for broad measles surveillance, alongside strong support for surveillance of infectious threats, aligns with the National Health Act [19] and the Disaster Management Act [20]. These Acts legally empower health bodies to bypass individual consent to mitigate severe epidemiological threats. This public intuition is further backed by major South African case law. In the landmark case *Minister of Health of the Western Cape v Goliath* [25], the court ruled that individual rights (such as privacy and freedom of movement) must be legally restricted if an individual poses a severe and imminent communicable threat to the broader community. Further, in *Ngobeni* [26], the High Court explicitly ruled that the strong privacy interests of patients were legally outweighed by the urgent "public interest" of managing the fallout of a severe outbreak, legally permitting the NHLS to share health data and by implication, access to compensation allowable through class action, without standard individual consent. The respondents in our study implicitly agree with the High Court: when tracking dangerous outbreaks, community safety takes legal precedence over granular privacy concerns.

To successfully expand WES in South Africa, policy and practice must adapt in four key areas. First, public health authorities must launch targeted, plain-language education campaigns to improve basic awareness of WES and build long-term trust. Second, local political leaders, such as ward councillors, should be educated on WES governance boundaries so they can support public health initiatives without compromising health data privacy rules. Third, public health agencies must establish clear mechanisms for sharing surveillance results back with communities in transparent, non-stigmatizing ways. Finally, South African health policies should be strengthened to explicitly clarify the regulatory status of WES as a disease surveillance tool under the National Health Act, including clear principles for data governance, use, and public reporting. Although WES is already used in South Africa to monitor pathogens such as poliovirus, measles, and SARS-CoV-2, clearer governance is needed as its use expands.

## Limitations

The small, localized sample size (N = 100) limits the generalizability of our results to the broader South African population, particularly rural areas with different sanitation systems. Second, self-selection and convenience bias may have led individuals with extreme privacy concerns or deep institutional mistrust to opt out, meaning the identified "privacy-concerned" group may reflect moderate rather than extreme views. Third, translating complex technical concepts like WES into local vernacular created challenges in maintaining clear conceptual meaning, which may have caused misunderstandings among participants. Finally, because the study focused on measles, public support for WES may not apply to all biological and chemical health targets that could carry higher social stigma.

## CONCLUSION

This is the first study to combine public opinion with legal Acts, policy documents, and judicial rulings for a low- or middle-income country. Importantly, this study shows that public support for WES in South Africa. Acceptability depends on keeping data combined at a city-wide level (anonymity by aggregation) and placing governance in the hands of trusted health bodies. These public preferences align directly with South African privacy laws and health Acts. Establishing WES-specific legal frameworks, expanding plain-language public education, and maintaining strong privacy safeguards will enable South Africa to integrate WES into public health surveillance with appropriate and sufficient regard for bioethical and legal requirements.

## Declarations

### Ethics approval and consent to participate

This study was conducted in accordance with the Declaration of Helsinki. The study protocol was approved by the Human Research Ethics Committee (HREC) of the University of the Witwatersrand (Clearance Certificate No. M250305). All participants provided written informed consent prior to administration of the survey instrument. Participation was voluntary, and all responses were anonymized using unique numerical identifiers.

## Consent for publication

Not applicable. The manuscript does not contain any individual person’s data, personal identifiers, or direct clinical images.

## Availability of data and materials

The datasets generated and analysed during the current study are provided in the supplementary information.

## Authors’ contributions

RM, RHH, and KM conceptualized and designed the study. SM provided administrative support for all aspects of the study. RM led data collection, survey administration, data analysis, submission for ethics review, and drafted the initial manuscript. SM and LR supported data collection and survey administration. KM and MY obtained funding. RHH and KM provided overall study supervision and oversight, and critically revised the manuscript for important intellectual content. All authors read and approved the final manuscript.

## Acknowledgements

We extend special thanks to the National Institute for Communicable Diseases (NICD) wastewater sampling team for their invaluable assistance with questionnaire administration and technical field support. Finally, we express our sincere appreciation to all community members who participated in this study.

## Questionnaire

This questionnaire is intended to collect data on your perceptions regarding acceptance of use of wastewater for community health monitoring in South Africa.

□ □

### Demographics□

Mark only one circle (in the options section)

**2.** What gender do you identify as?□□

`o` Male□ □
`o` Female□ □
`o` Non-binary /third gender
`o` Other
`o` Prefer not to say
**2.** What is your age range?

`o` 18 – 24
`o` 25 – 34
`o` 35 – 44□ □
`o` 45 – 54□ □
`o` 55 – 64
`o` 65 – 74
`o` 75 or older
**3.** What is the highest level of education that you have completed?□□

`o` No formal education
`o` Primary school
`o` Some secondary school
`o` Matric
`o` Tertiary qualification
**4.** What type of toilet do you use at home?

`o` No toilet – I use the bush
`o` Pit latrine
`o` Flush toilet to a septic tank (A septic tank is an underground system that collects and treats wastewater from your home. You would have had to have paid for this installation)
`o` Flush toilet connected to a sewage system (A flush toilet connected to a sewage is a toilet where waste is flushed away through pipes to a large underground system that safely carries it to a treatment plant for cleaning)

### Block A□□

Wastewater surveillance means taking samples from sewage and testing this water in the laboratory. Results will help the health department to know if illnesses such as measles are present in the community. Then the health department can act quickly to prevent the illness from spreading.

Mark only one option below

**5.** Have you heard, seen or been told about wastewater monitoring before?

`o` Yes
`o` No
`o` I don’t know
6. The municipality conducts environmental activities to protect the public’s well-being. Would you be comfortable with monitoring wastewater for the following items? For each activity, tick the most appropriate option below to indicate your level of comfort. □ □□ □

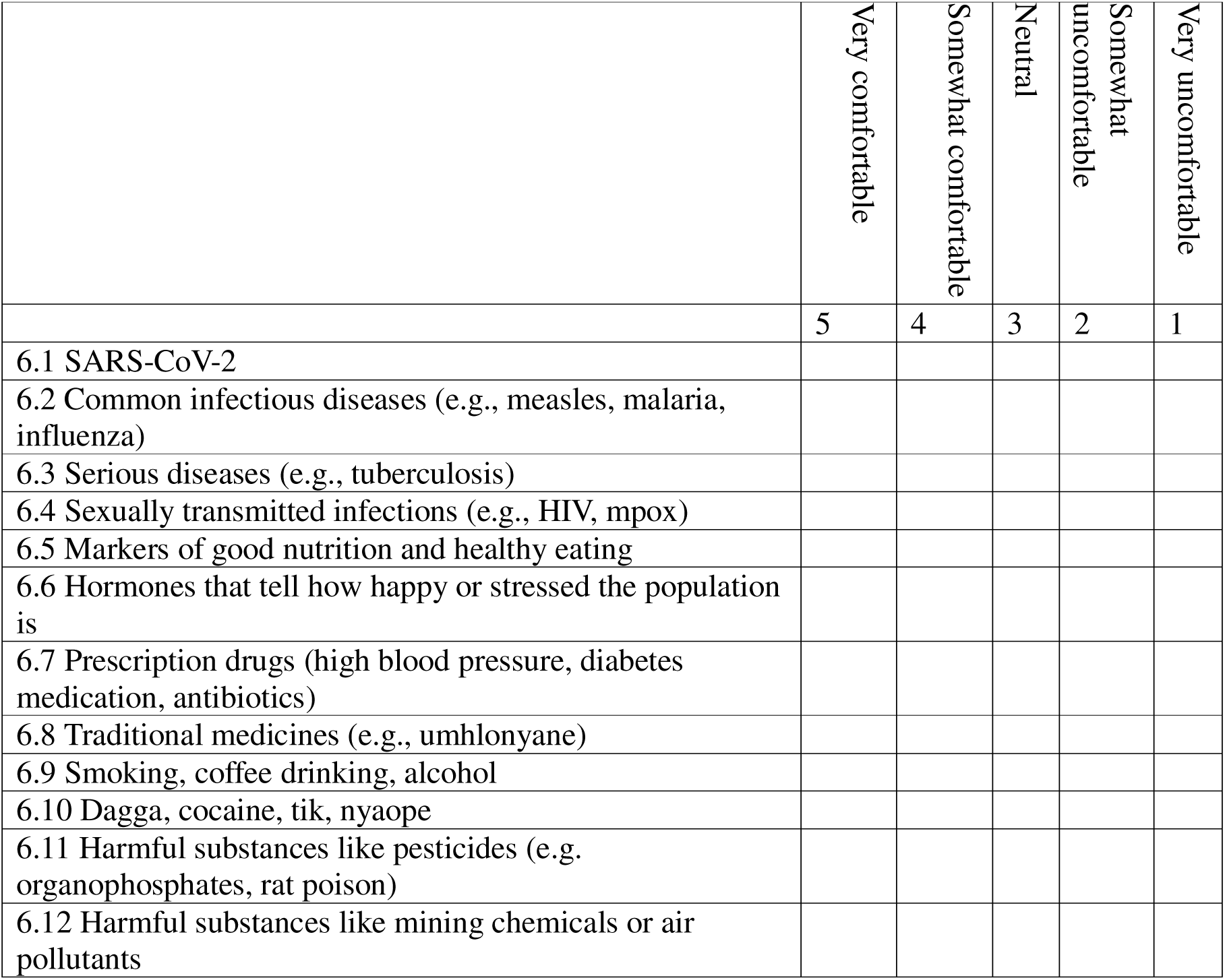

### Block B□□

7. Now that you know we can monitor wastewater, in which types of places do you think wastewater monitoring should be conducted? Please select all that apply □

`o` The entire city □
`o` This suburb/community
`o` Businesses/ factories
`o` Shopping centres/ taxi ranks
`o` Schools, Colleges and Universities
`o` Prisons□ □
`o` Informal settlements
`o` home
`o` Church/mosque/synagogue
`o` I would not support any monitoring of wastewater
**8.** Do you feel there are any areas where wastewater should not be monitored? **Mark all that apply.**□ □

`o` The entire city □
`o` This suburb/community
`o` Businesses/ factories
`o` Shopping centres/ taxi ranks
`o` Schools, Colleges and Universities
`o` Prisons □ □
`o` Informal settlements
`o` homes
`o` Church/mosque/synagogue
`o` I support any monitoring of wastewater
**9.** We want to find out what your thoughts are around wastewater surveillance for measles, a highly contagious disease caused by a virus. So, for each statement, tick the most appropriate option to show how much you agree with the following statements.

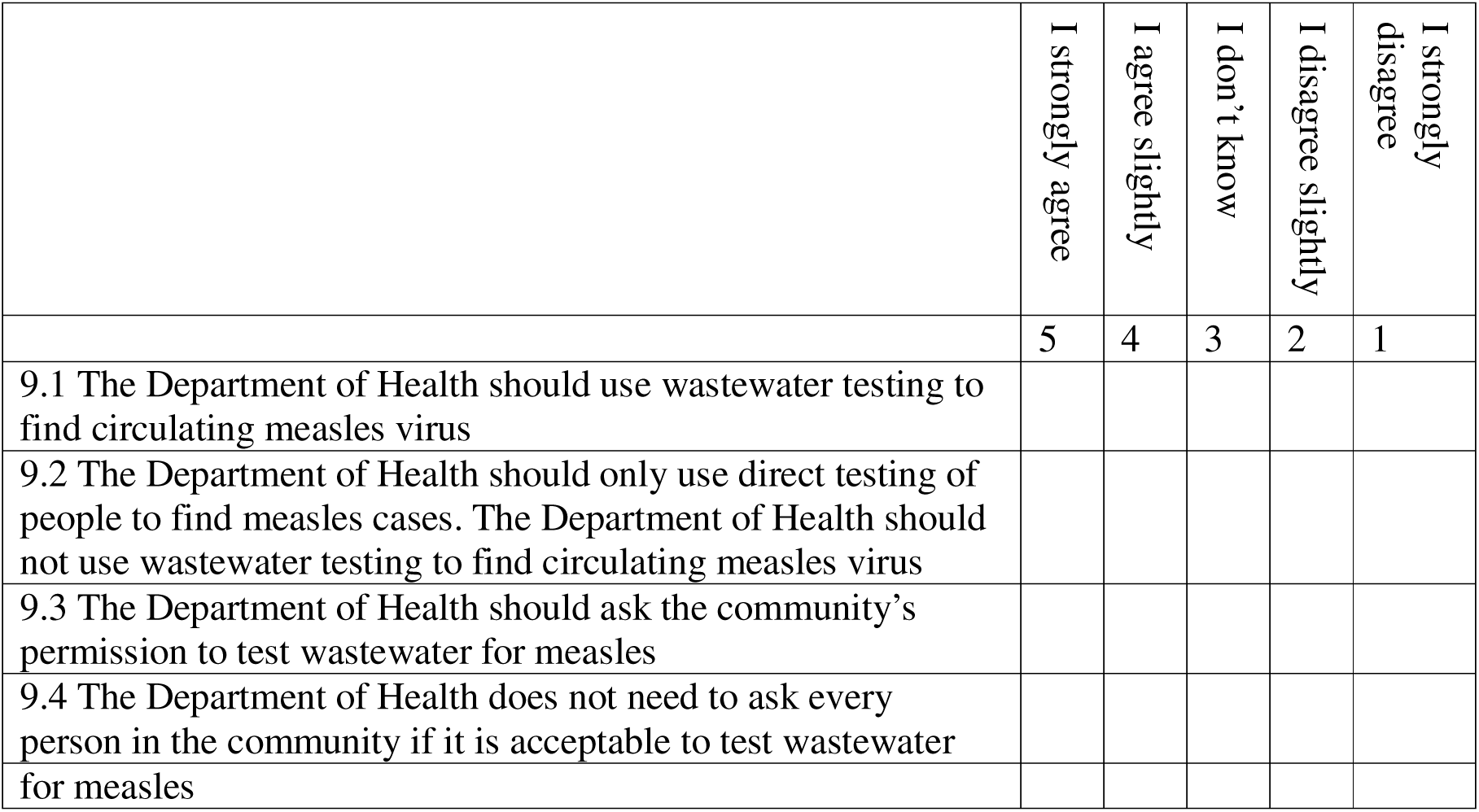

□

**10.** In your own opinion, can wastewater testing for measles identify a single case of measles in the community? Do you believe that wastewater testing can be traced back to identify individual contributors?

`o` Yes
`o` No
`o` I don’t know

### Block C□□

11. Do you consider wastewater monitoring to be an invasion of privacy?□□Mark one option below

`o` Yes
`o` No
`o` I don’t know
How comfortable are you sharing your personal health information (for example your blood test results, medications, vaccination records) with the following groups?□ □

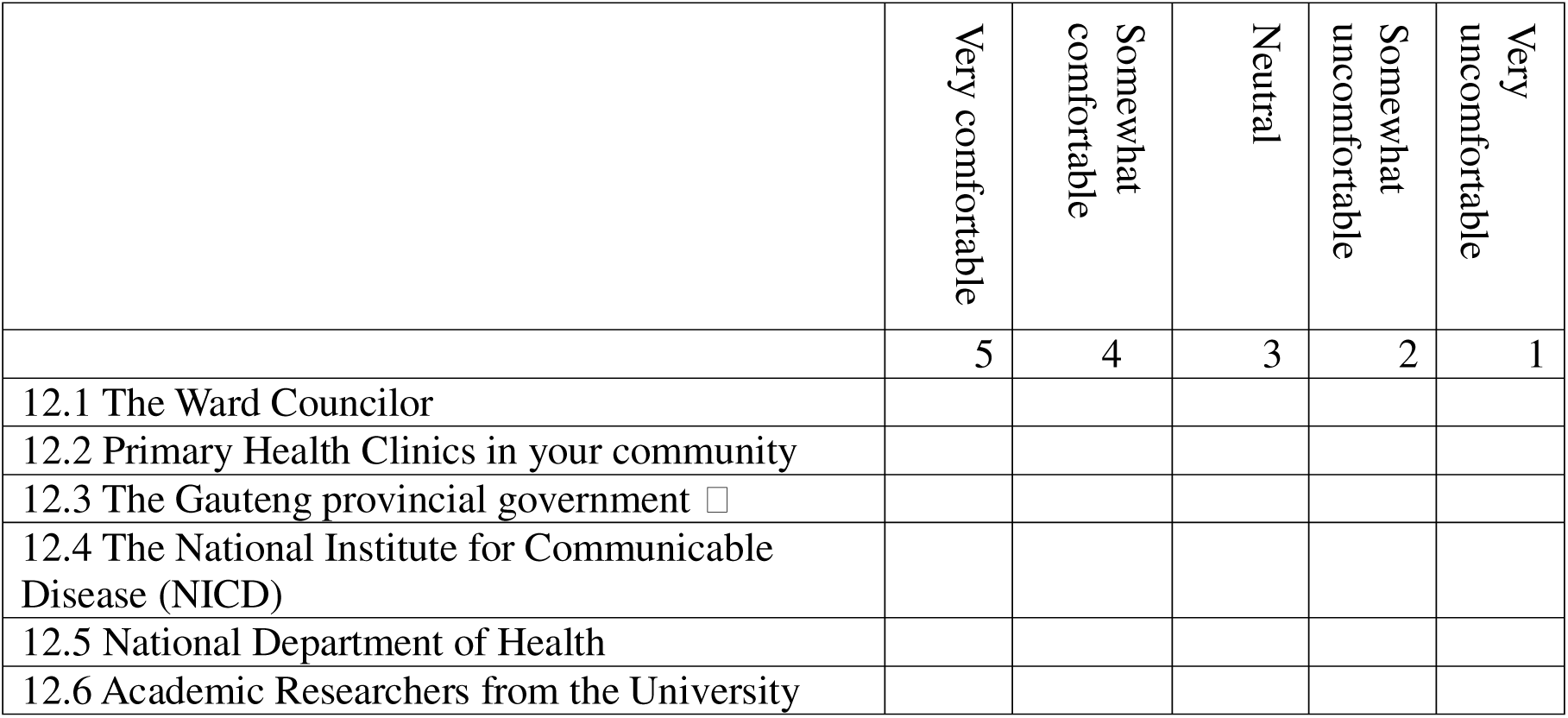

12. How comfortable are you for the Department of Health to share information they get from wastewater collected in your community with the following groups?□□Use the corresponding number(s) below to indicate your level of support or opposition□

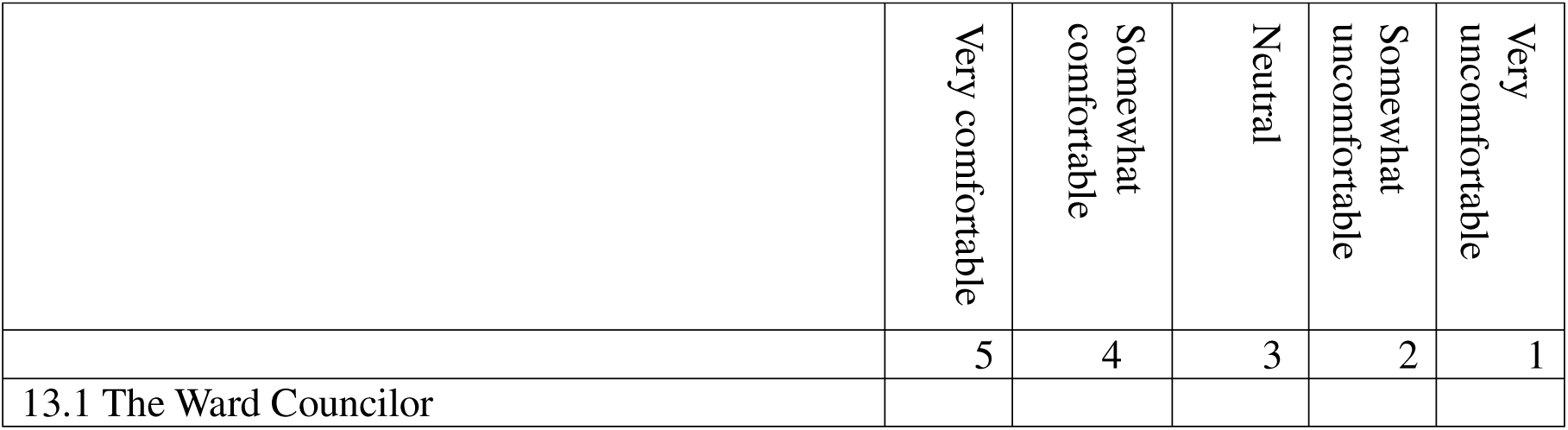
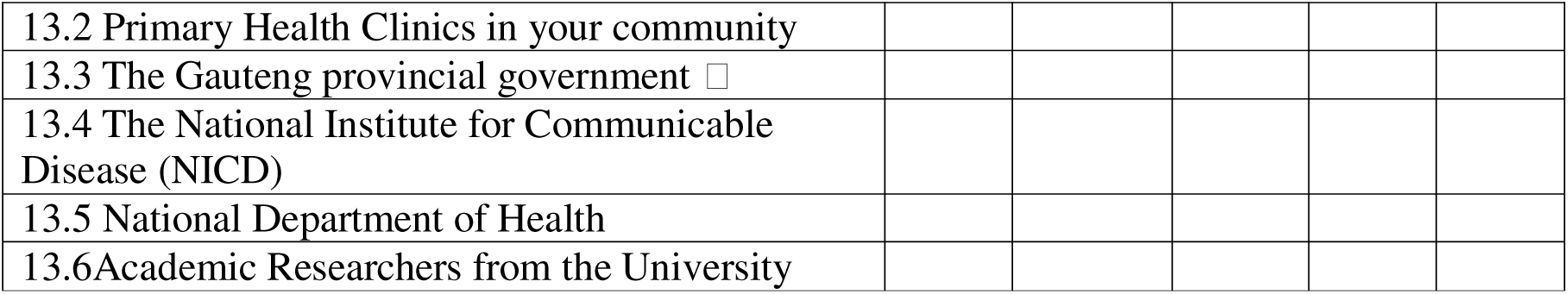

13. How much of your information are you comfortable sharing? Use a number to show your level of willingness

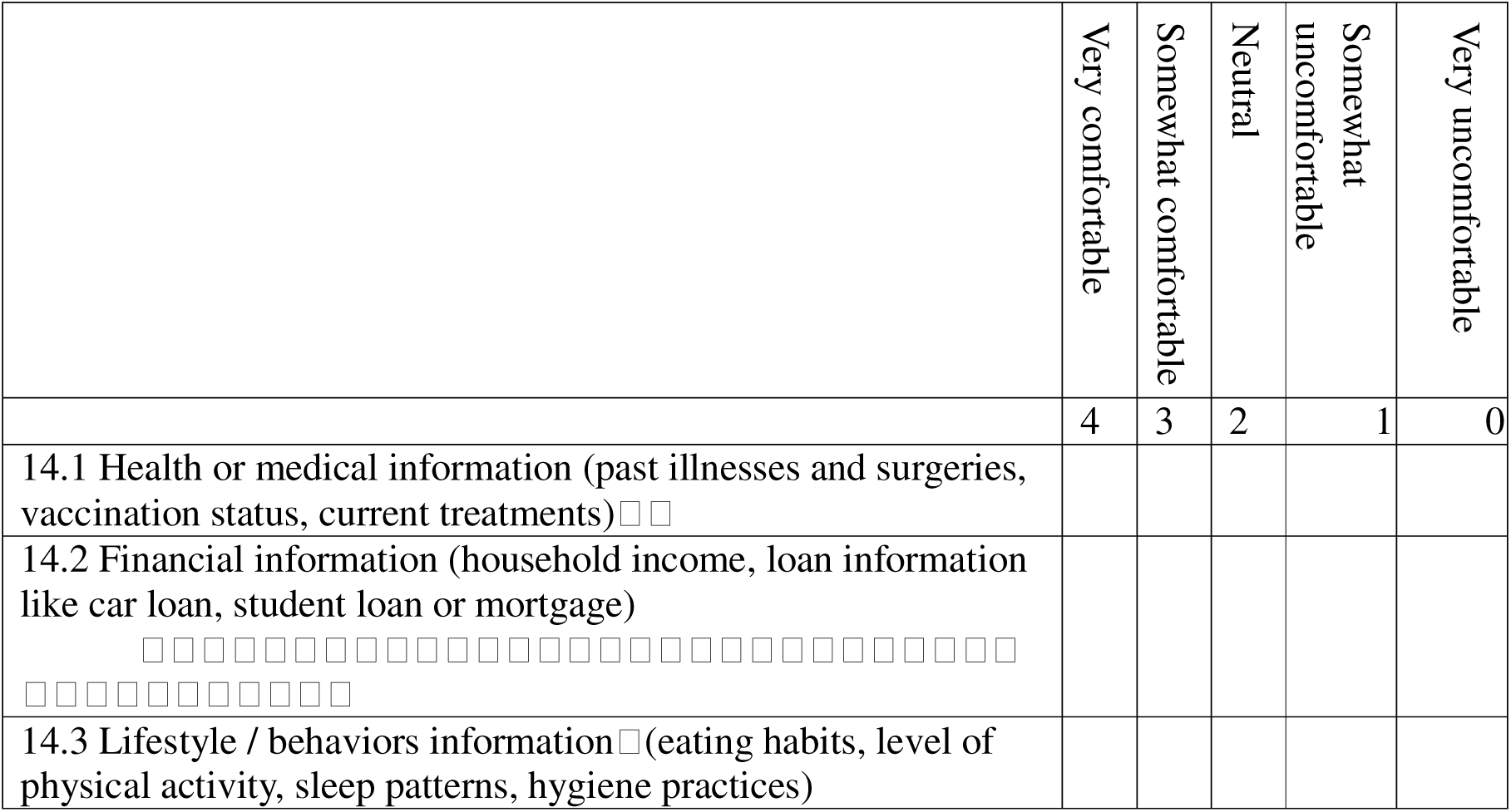

### Block D

14. Do you have any concerns about samples being taken from flush toilets in this area?□□Use the block below to write any concerns

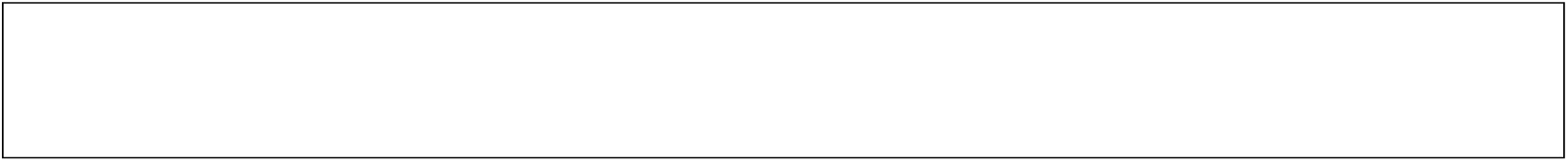

## Summary Demographics

Demographic attributes, sewer connections and WES awareness amongst 100 respondents of a wastewater ‘knowledge attitudes and practice’ questionnaire administered between 5–27 November 2025 in three Metros of Gauteng province.

Number of participants (n)

|  | Number of participants (n) |  |  |  |
| --- | --- | --- | --- | --- |
|  | CoE (79) | CoT (6) | CoJ (15) | All (100) |
| **Gender (n, % male)** | 48 (60%) | 3 (50%) | 12 (80%) | 63 |
| **Gender (n, % female)** | 31 (39%) | 3(50%) | 3 (20%) | 37 |
| **Age category (n, %)** |  |  |  |  |
| 18-24 | 9 (11%) | 3 (50%) | 3 (20%) | 15 |
| 25-34 | 34 (43%) | 2 (33%) | 7 (46%) | 43 |
| 35-44 | 25 (31%) | 0 | 4 (26%) | 29 |
| 45-54 | 5 (6%) | 1(17%) | 0 | 6 |
| 55-64 | 5(6%) | 0 | 1(7%) | 6 |
| 65 - 74 | 1(1%) | 0 | 0 | 1 |
| **Highest education level (n,%)** |  |  |  |  |
| No formal education | 2 (2.53%) | 0 | 0 | 2 |
| Primary School | 6 (8%) | 1(17%) | 1 (6%) | 8 |
| Some secondary school | 11 (14%) | 0 | 6 (40%) | 17 |
| Matric | 33 (42%) | 5 (83%) | 8 (53%) | 46 |
| Tertiary | 27 (34.17) | 0 | 0 | 27 |
| **What kind of toilet do you use (n,%)** |  |  |  |  |
| No toilet | 3(4%) | 0 | 0 | 3 |
| Pit | 4(5%) | 0 | 6(40%) | 10 |
| Flush connected to septic | 18 (23%) | 4 (67%) | 6 (40%) | 28 |
| Flush connected to sewer | 54 (68%) | 2 (33%) | 3(20%) | 59 |

\*CoE = City of Ekurhuleni; CoT = City of Tshwane; CoJ = City of Johannesburg

